# Pleural Empyema in Tijuana, Mexico, and the impact of Pneumococcal Conjugate Vaccines: Twenty Years of Active vs. Passive Surveillance Incorporating PCR - Evidence Supporting the Need for Enhanced Active Monitoring and Molecular Diagnosis

**DOI:** 10.64898/2025.12.26.25342828

**Authors:** Erika Z. Lopatynsky-Reyes, Jaime A. Rodriguez-Valencia, Enrique Chacon-Cruz

## Abstract

In Tijuana General Hospital, Mexico, pleural empyema (PE) was monitored through passive surveillance (2000 - 2005), active surveillance with PCR (2005 - 2018), and again through passive surveillance (2022 - 2024). The implementation of active surveillance combined with molecular diagnostics markedly improved pathogen detection and enabled assessment of the 7-valent (PCV7, 2005 - 2012) and 13-valent (PCV13, 2012 - 2018) pneumococcal conjugate vaccines. This strategy yielded precise insights into the etiology of PE in children and adolescents, enhanced antimicrobial susceptibility profiling to guide antibiotic therapy, and provided strong evidence supporting the high effectiveness of PCV13. This study also provides insights towards promoting evidence-based decision-making towards effectiveness and safety outcomes of vaccines and potential new implementations to the national immunization program.

The thirteen years of active surveillance were approved by the Institutional Review Board (IRB) of Hospital General de Tijuana. Because one of the objectives of this study was to compare passive and active surveillance, the use of data obtained from hospital records for passive surveillance was also reviewed and approved by the IRB. Patient identities were never disclosed, and no biological samples were retained for future analyses.

## Introduction

Pleural empyema (PE) is a serious complication of bacterial pneumonia (BN) associated with high morbidity, prolonged hospitalization, potential sequelae, and, in some cases, death [1-3]. Among children beyond the neonatal period, the leading causative pathogens of both BN and PE are *Streptococcus pneumoniae, Staphylococcus aureus*, and *Haemophilus influenzae* [3-6].

At Tijuana General Hospital, Mexico, pediatric PE was monitored across three distinct phases: passive surveillance (2000–2005), active surveillance incorporating polymerase chain reaction (PCR) diagnostics (2005–2018), and a subsequent return to passive surveillance (2022–2024).

To date, no published studies have systematically compared passive and active surveillance for pleural empyema (PE). The objective of this study is precisely to evaluate the impact of implementing active surveillance by comparing the detection rates and characterization of all variables associated with PE during periods of passive versus active surveillance, as well as after active surveillance was discontinued.

## Methods

Pleural empyema (PE) was defined as the presence of purulent pleural fluid. All children aged 3 months to under 16 years with PE admitted to Tijuana General Hospital, Mexico, were included in the study. All data were compiled in a Microsoft Excel database, and descriptive statistical analyses were performed.

Three distinct surveillance periods were evaluated:

### 1. 2000–2005: Retrospective passive surveillance

Data were collected from hospital records of patients younger than 16 years diagnosed with PE.

### 2. 2005–2018: Prospective active surveillance

Active, prospective surveillance was initiated in the emergency department. Following pleural puncture, pleural fluid (PF) samples were immediately inoculated into radiometric broth media (BACTEC^®^, Becton Dickinson, Franklin Lakes, NJ, USA) and incubated at 37 °C with 5% CO. Final bacterial identification was performed using the Vitek/Microscan Vitek2^®^ system (bioMérieux, Hazelwood, MO, USA).

For real-time polymerase chain reaction (RT-PCR), DNA was extracted from PF using the QIAamp DNA Blood Mini Kit (QIAGEN^®^, Shanghai, China) according to the manufacturer’s instructions. A 200 μL PF aliquot was processed, and DNA was eluted in 100 μL of TE buffer. RT-PCR was performed on the Mx3000P qPCR System (Stratagene^®^, La Jolla, CA, USA), targeting seven common bacterial pathogens associated with PE via specific gene markers: *16S* (*E. coli*), *femA* (*S. aureus*), *hly* (*L. monocytogenes*), *ctrA* (*N. meningitidis*), *lytA* (*S. pneumoniae*), *bexA* (*H. influenzae*), and *cfb* (Group B *Streptococcus*).

*S. pneumoniae* serotypes were identified by Quellung reaction (Statens Serum Institut^®^, Copenhagen, Denmark) and/or multiplex PCR following the CDC-recommended sequential method. Serotype-specific primers were validated with individual reference strains and subsequently tested against 5–10 additional clinical isolates to ensure broad detection within each serotype. For *S. aureus*, methicillin resistance was determined using 6 μg/mL oxacillin in Mueller–Hinton agar supplemented with 4% NaCl, or by PCR detection of the *mecA* gene.

### 3. 2022–2024: Passive surveillance reinstatement

Following the COVID-19 pandemic (2020–2022), when hospital admissions were limited to SARS-CoV-2 cases, passive surveillance for pediatric PE was reestablished.

## Results

### 2000–2005: Retrospective passive surveillance

Between 2000 and 2005, 13 cases of PE were reported (2.6 cases per year). Pathogens were isolated by conventional culture in only 4 cases (30.7%): two *S. pneumoniae*, one *S. aureus*, and one *S. pyogenes*. Antimicrobial susceptibility testing was not performed for *S. pneumoniae* isolates. For *S. aureus* and *S. pyogenes*, testing by the disk diffusion method showed that the *S. aureus* strain was susceptible to oxacillin and glycopeptides, while the *S. pyogenes* strain was susceptible to penicillin.

### 2005–2018: Prospective active surveillance

Between 2005 and 2018, 64 cases of PE were actively identified (4.9 cases per year), representing a 1.9-fold increase compared with the 2000–2005 period (Figure 1). Bacterial pathogens were identified in 51 cases (79.7%), a 2.6-fold improvement in detection compared to the earlier period (Figure 2). The predominant organisms were *S*.*pneumoniae* (29; 56.8%), *S. aureus* (14; 27.4%), and *S. pyogenes* (3; 5.9%). Single cases were also attributed to *Peptostreptococcus sp*., *P. aeruginosa, K. oxytoca, S. salivarius*, and *S. milleri*.

**Figure 1.**
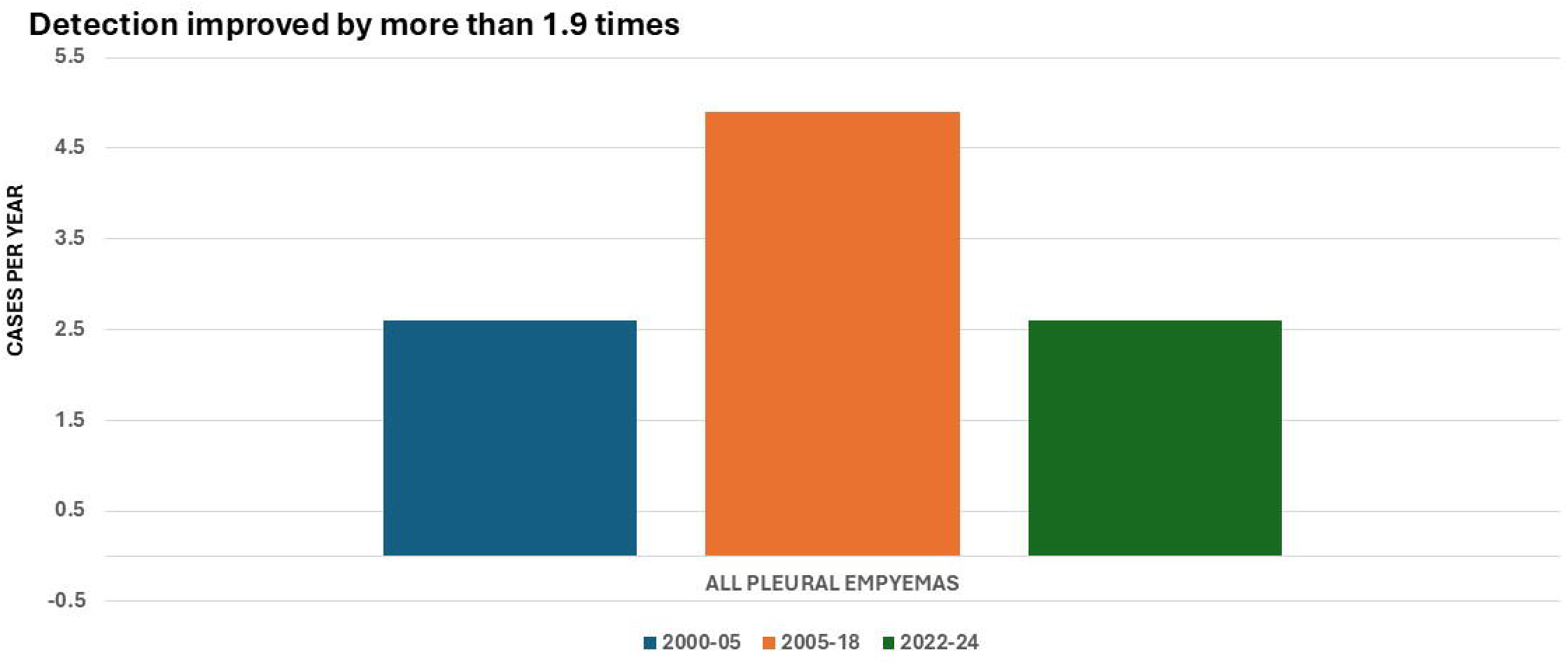
Annual Cases of all Pleural Empyemas in Children and Adolescents in Tijuana, Mexico: Active Surveillance + PCR (2005-2018) vs. Passive Surveillance (2000-2005, 2022-2024)

**Figure 2.**
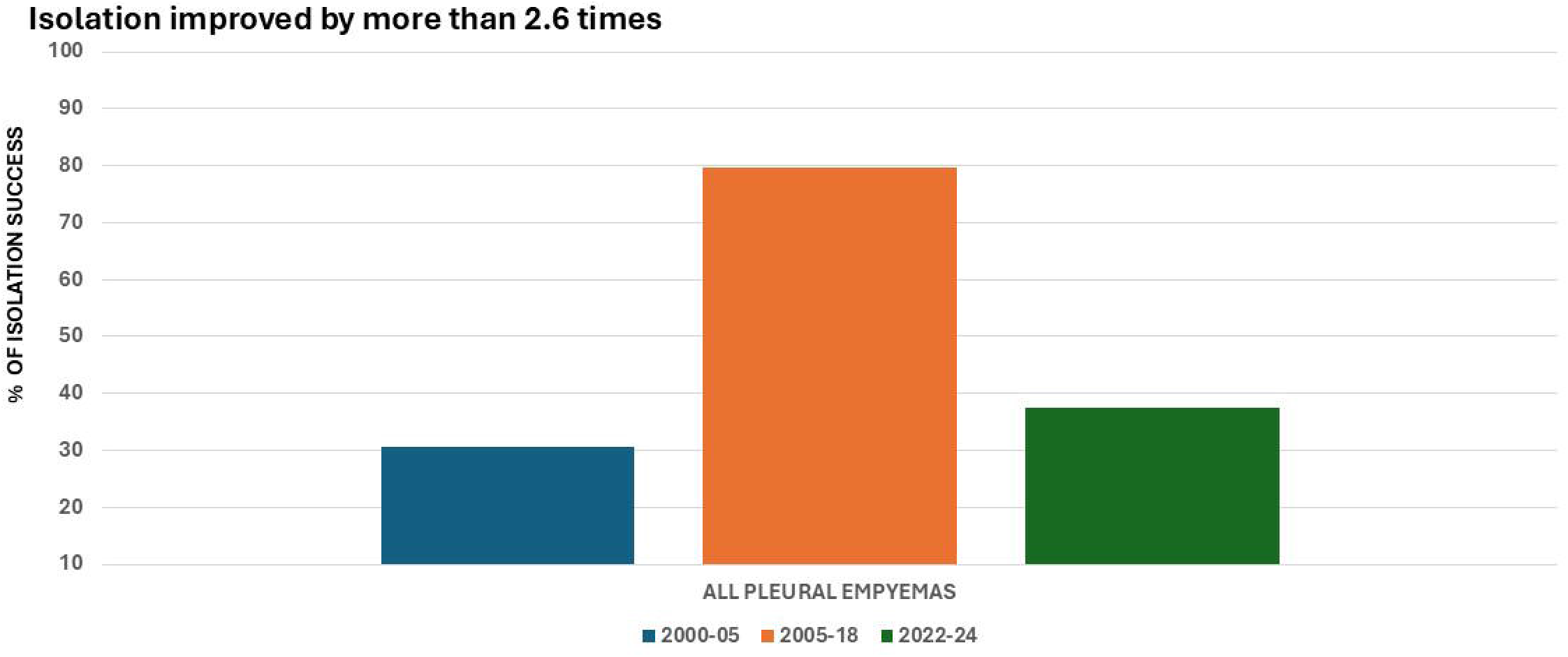
Bacterial Isolation (%) of Pleural Empyemas in Children and Adolescents in Tijuana, Mexico: Active Surveillance + PCR (2005-2018) vs. Passive Surveillance (2000-2005, 2022-2024)

Pneumococcal PE: During the PCV7 implementation period (2005–2012), a reduction in overall PE cases was observed; however, a resurgence occurred from 2009 onward, primarily driven by pneumococcal serotypes 19A, 3, 14, and 7F. Following the introduction of PCV13 in 2013, there was a 53.8% relative reduction in all pneumococcal PE cases, including an apparent elimination of serotype 19A and an 81.8% decrease across all vaccine-covered serotypes, with minimal emergence of non-vaccine serotypes.

Vaccine Effectiveness and Pathogen Shift: The PCV13 vaccine demonstrated an 81.8% effectiveness against pneumococcal PE caused by PCV13-included serotypes, and a 53.8% reduction in overall pneumococcal PE irrespective of serotype (Figure 3). However, no net reduction was observed for all-cause PE, driven by a rise in *S. aureus*-associated cases during the final three years of active surveillance (13 cases post-PCV13 implementation, Figure 3). Notably, all but one *S. aureus* isolated from pleural fluid were methicillin-resistant (MRSA), including one strain exhibiting inducible clindamycin resistance (D-test positive).

**Figure 3.**
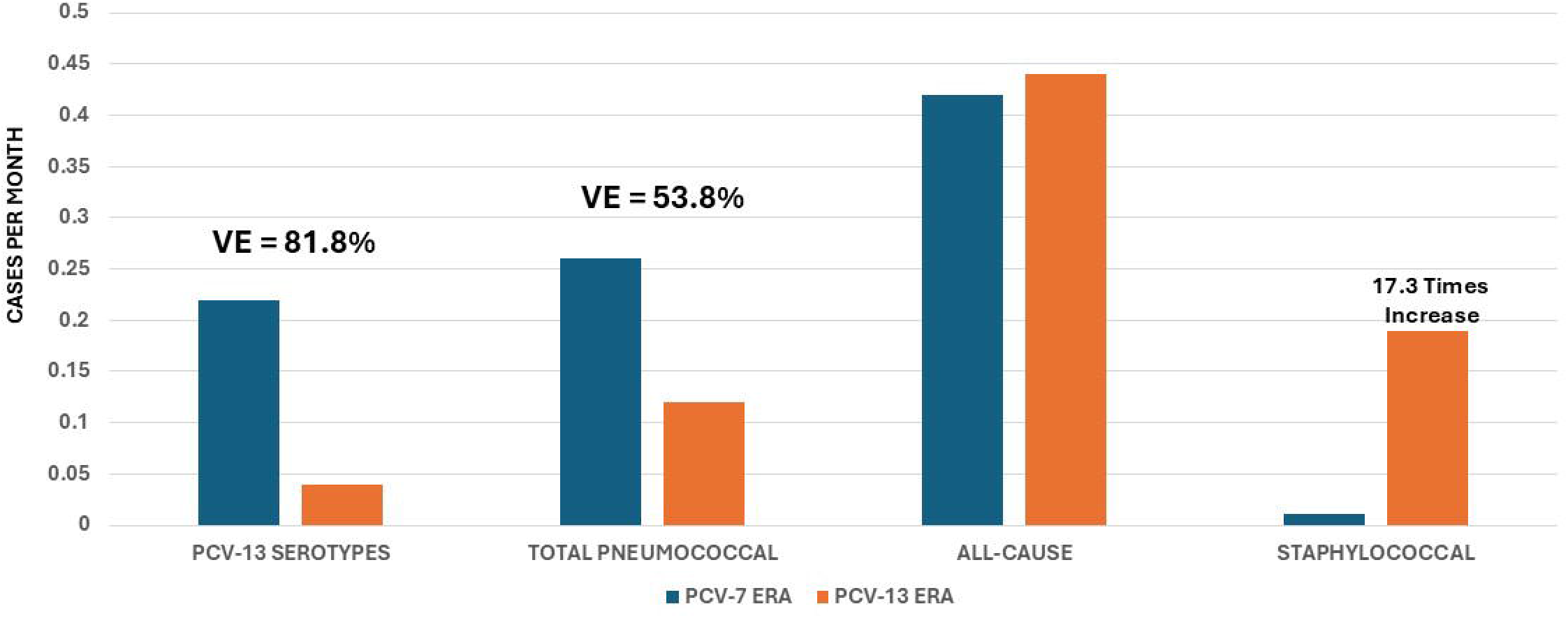
Impact of PCV13 vs. PCV7 on Pleural Empyema: PCV-13 Vaccine Serotypes, Total Pneumococcal, All-Cause, and Staphylococcal Cases.

Clinical, laboratory, tomographic, treatment, and outcome data for each patient are already published and available in reference [7].

### 2022–2024: Passive surveillance reinstatement

Between 2022 and 2024, eight cases of PE were reported (2.6 cases per year), representing a 1.9-fold decrease compared with the 2005–2018 period (Figure 1). Pathogens were isolated in only three cases (37.5%), reflecting a 2.1-fold reduction in bacterial identification relative to the prior period (Figure 2). The isolated organisms were *S*.*pneumoniae, S. aureus*, and *K. pneumoniae*—one of each—with no antimicrobial susceptibility data available.

## Discussion

As mentioned in the introduction, the objective of this study is precisely to evaluate the impact of implementing active surveillance by comparing the detection rates and characterization of all variables associated with PE during periods of passive versus active surveillance, as well as after active surveillance was discontinued.

For pneumococcal disease, multiple studies have demonstrated that active surveillance combined with molecular diagnostics (PCR) is crucial—not only for improving case detection across the spectrum of pneumococcal infections, but also for monitoring serotype dynamics following the introduction of pneumococcal conjugate vaccines (PCVs). This approach enables a more accurate characterization of clinical outcomes, sequelae, morbidity, mortality, and the identification of other pathogens involved [8-15].

Particularly for PE, there are various publications of pediatric PE by implementing active surveillance:

In Germany, Forster *et al*. conducted a prospective study using PCR to evaluate the impact of prehospital antibiotic therapy on clinical outcomes and pathogen detection in 1,724 children with parapneumonic pleural effusion or empyema between 2010 and 2018. Their findings showed that while prehospital antibiotic administration significantly reduced bacterial pathogen detection by culture, it did not affect detection by PCR. Moreover, prehospital antibiotic use was associated with fewer infectious complications but did not influence the overall duration of illness [16].

In Turkey, pleural fluid samples from 156 children diagnosed with pneumonia complicated by empyema across 13 hospitals between 2010 and 2012 were collected via thoracentesis and tested for 14 *S. pneumoniae* serotypes/serogroups using a Bio-Plex multiplex antigen detection assay. The estimated serotype coverages for PCV7, PCV10, and PCV13 were 16.3%, 45.4%, and 60%, respectively. The authors concluded that active surveillance studies are essential to monitor shifts in *S. pneumoniae* serotypes causing empyema, in order to guide optimal pneumococcal vaccine selection [17].

Similar results were found in children with parapneumonic pneumonia in Italy and Japan [18, 19].

Our first publication in 2019 not only demonstrated an increase in the detection of pleural empyema (PE) cases but also provided enhanced insights into: (1) clinical, demographic, and patient outcomes; (2) the impact of PCV7 and PCV13 on pneumococcal serotype distribution and replacement; and (3) the contribution of pathogens other than *Streptococcus pneumoniae* [7].

Additionally, through active surveillance combined with molecular diagnostics (PCR), we observed the emergence of PE cases caused by *S. aureus*.

Although studies from the United States have not shown an increase in *S. aureus*–associated pneumonia following PCV13 implementation [20], several reports have documented a potential rise in *S. aureus*–related PE cases after the introduction of PCV13:

In Italy, Carloni *et al*. analyzed 43 pediatric cases and found a significant reduction in hospitalizations for necrotizing pneumonia (including pleural empyema) between the early (1.5/1000 admissions/year) and intermediate (0.35/1000 admissions/year) post-PCV13 periods (*p* =.001), followed by an increasing trend thereafter. *S. pneumoniae* remained the predominant pathogen in both periods (pre-PCV13: 11/18, 61%; post-PCV13: 13/25, 52%), with serotype 3 being the most frequent strain (pre-PCV13: 3/11, 27%; post-PCV13: 4/13, 31%). Although the overall etiology did not change substantially over time, most infections caused by *S. pyogenes* or *S. aureus* occurred during the post-PCV13 period [21].

In Germany, Lise *et al*. conducted a prospective study incorporating PCR in 1,447 children with parapneumonic pleural effusion or empyema between 2010 and 2017. Their findings showed an increase in *S. pyogenes* infections following PCV13 introduction, but no corresponding rise in *S. aureus* cases [22].

Our study is the first to directly compare the benefits of implementing active surveillance and to demonstrate the substantial loss of essential epidemiologic information that occurs when this methodology is discontinued.

Moreover, beyond pathogen identification and shifting serotype patterns, this study underscores that the true value of active surveillance lies in its ability to drive evidence-based decision making for national immunization programs by complementing passive surveillance and clinical trials [22]. Robust surveillance systems—especially those strengthened with enhanced and molecular tools such as genotyping—generate the high-quality, timely, and comprehensive data needed to evaluate vaccine effectiveness, ensure safety, sustain public confidence, and guide rational vaccine introduction [23-26]. Importantly, these systems create a continuous quality-improvement loop, enabling health authorities to monitor disease epidemiology, rapidly detect vaccine-preventable threats, and refine immunization strategies as new evidence emerges. In this way, active surveillance becomes not only a technical activity, but a foundational mechanism that informs clinical practice, shapes public health policy, and ultimately strengthens both routine immunization efforts and future pandemic preparedness [27-28].

## Conclusions

Active surveillance, incorporating immediate culturing and PCR sampling in the emergency department, doubled the detection rate of pleural empyema cases compared to passive surveillance. The combination of rapid culture and PCR enabled a 2.6-fold increase in bacterial identification, improved antimicrobial susceptibility testing, and supported more informed antibiotic decision-making.

Establishing active surveillance with molecular diagnostics also enabled evaluation of pneumococcal conjugate vaccine effectiveness, with PCV13 demonstrating a strong impact in reducing pneumococcal pleural empyema, particularly those caused by vaccine-included serotypes.

As a surprising finding, PCV13 effectiveness for all pneumococcal PE was also associated with an increase on staphylococcal PE. Reinstating active surveillance with PCR is strongly recommended to ensure comprehensive pathogen detection and accurate epidemiologic monitoring and drive evidence-based vaccine policy-decision-making.

## Data Availability

All data produced in the present study are available upon reasonable request to the authors

